# Mapping Psilocybin-Assisted Therapies: A Scoping Review

**DOI:** 10.1101/2019.12.04.19013896

**Authors:** Ron Shore, Paul Ioudovski, Sandra McKeown, Eric Dumont, Craig Goldie

## Abstract

We conducted a scoping review on psilocybin-assisted therapy for addiction, depression, anxiety and post-traumatic stress disorder. Psilocybin is a naturally-occurring tryptophan derivative found in species of mushroom with psycho-active properties. From 2022 records identified by database searching, 40 publications were included in the qualitative synthesis from which we identified 9 clinical trials with a total of 169 participants. Trials used a peak-psychedelic model of therapy, emphasizing inward journey through the use of eyeshades, set musical scores and with medium to high doses of psilocybin. No serious adverse effects were reported; mild adverse effects included transient anxiety, nausea and post-treatment headaches. Overall, the 9 trials all demonstrated safety, tolerability and preliminary efficacy in the treatments of obsessive-compulsive disorder, substance use disorder, treatment-resistant unipolar depression, anxiety or depression in patients with life-threatening cancer and demoralization among long-term AIDS survivors.The literature was found to be early and exploratory, with several limitations: only 5 were randomized controlled trials, small and homogenous patient sample size, difficulties in blinding, and the confounding influence of psychological supports provided. Further research is indicated to establish effectiveness for these and other indications, with a more diverse range of patients, and with differing program and dosing modalities.

## Introduction

Psilocybin (4-phosphoryloxy-*N,N*-dimethyltryptamine) is a naturally-occurring tryptamine indolealkylamine and a prodrug to psilocin (4-Hydroxy-N,N-dimethyltryptamine), a central serotonin 5HT2A receptor agonist. Along with Lysergic acid diethylamide (LSD) and N, N-Dimethyltryptamine (DMT), psilocybin is considered a classic serotonergic hallucinogen, psychedelic. First isolated and identified in 1958 by Albert Hoffman from the strain *Psilocybe Mexicana* (Hoffman, 1958), psilocybin is the main psychoactive molecule found in “magic mushrooms”. Sacramental use of the Psilocybe genus of gilled fungi in Mesoamerica has been dated to 500 BC (Guerra-Doce, 2015), with shamanic ceremonial traditions documented among many indigenous groups throughout central-southern Mexico, as well as among the Yurimagua of Peru (Schultes et al., 1992, Guzman, 2008, McKenna & Riba, 2015)

Onset of action for psilocybin is 20-40 minutes after oral ingestion, with peak levels experienced after 60–90 minutes; the overall duration is 4–6 hours for oral administration (Tylša et al., 2013). Psilocin is largely excreted after 3 hours, and completely eliminated after 24; half-life is 2.5 hours. (Hasler et al., 2002; Tylša et al., 2013). Psilocybin has a very high safety ratio and a low risk profile even in unsupervised, naturalistic settings (Schenberg, 2018, Nutt et al., 2010).

Serotonin receptors regulate a range of processes including learning and memory, sleep and wake cycles, thermoregulation, appetite, sexual behaviour, pain, motor activity and aspects of autonomic function (Flanagan & Nichols, 2018). Nichols identifies the 5-HT2AR, principally expressed in the cortex (Pazos et al., 1987) as the key site of action, conjecturing that the novel patterns of global neurological interconnectivity produced by psilocybin, when combined with the appropriate clinical preparation and setting, can result in a subsequent, beneficial “rewiring” of brain networks away from previous pathological patterns and more similar to pre-disease states. (Nichols et al., 2017).

Recent investigations have indicated that the phenotypic expression of the 5-HT2AR genotypes is bound to and dependent on environmental context (Jokela et al. 2007), and this sensitivity to setting is integral to the functioning of 5-HT2A signalling (Carhart-Harris & Nutt, 2017). Psychedelic therapy has long paid heed to the importance of both set — personality, mindset, affect, expectations (Metzner and Leary, 1967) and setting (Leary, Litwin & Metzner R. 1963, Hartogsohn, 2017). The triad of drug, set and setting is foundational to understanding the total drug effect of any psycho-active substance (Zinberg, 1984) and central to harm reduction (McElrath and McEvoy, 2002; Shewan et al., 2000).

Previous reviews have been published summarizing research about the clinical potential of serotonergic psychedelics (Dos Santos 2016, Reiche et al. 2018, Schenberg 2018, Studeris 2011), including psilocybin (Johnson and Griffiths, 2017). We conducted a scoping review to identify, summarize and map the literature on psilocybin-assisted therapeutic trials. Scoping reviews, guided by an established, rigorous and replicable methodology (Arksey and O’Malley 2005) are used to map the nature, features and volume of existing literature in a given field, helping to clarify existing working definitions and conceptual boundaries. (Peters, 2015).

Scoping reviews provide a basis in evidence for the evolution of both research and clinical practice, in this case by understanding the patients selected, the interventions used, and the outcomes noted for psilocybin-assisted therapies. Our scoping review was guided by the research question, *“what are the treatment variables and outcomes associated with psilocybin-assisted therapy*?”

## Methods

This scoping review search was specific to: anxiety, depression, substance use disorder and post-traumatic stress disorder. Variables included both patient and intervention characteristics. Outcomes included any documented changes in health status and both positive and negative outcomes noted.

A comprehensive search approach (Appendix 1) was used to locate published and unpublished studies. A preliminary search was conducted in Ovid MEDLINE using a combination of keywords and subject headings, followed by an analysis of relevant citations to identify additional relevant keywords and subject headings. A refined search using all identified keywords and subject headings was then executed in Ovid MEDLINE (1946 onward) and translated in the following resources: Ovid Embase (1947 onward), PsycINFO (1806 onward), EBM Reviews: Cochrane Central Register of Controlled Trials (1991-present), Web of Science Core Collection (1900 onward), and ProQuest Dissertations and Theses (1861 onward). Database searches were conducted in December 2018 and updated in October 2019; no language or date restrictions were applied though non-English texts would eventually be excluded. Finally, the reference lists of all eligible reports and articles were hand-searched to identify any additional studies.

From records identified through database screening and using Covidence online software, duplications were removed, records screened, assessed for eligibility and included or excluded from full-text review (Table 1.) Inclusion criteria were: psilocybin, therapy/treatment, outcomes measured. Exclusion criteria were: duplicate publication, not research study, no outcomes measured, no therapeutic goal, not about psilocybin, naturalistic/recreational use, or microdosing. Reviewers read the full text of selected studies and each extracted data using an original data extraction tool. Reviewers cross-checked for homogenous process quality control and consensus was necessary when conflicts arose.

**Table 1.**
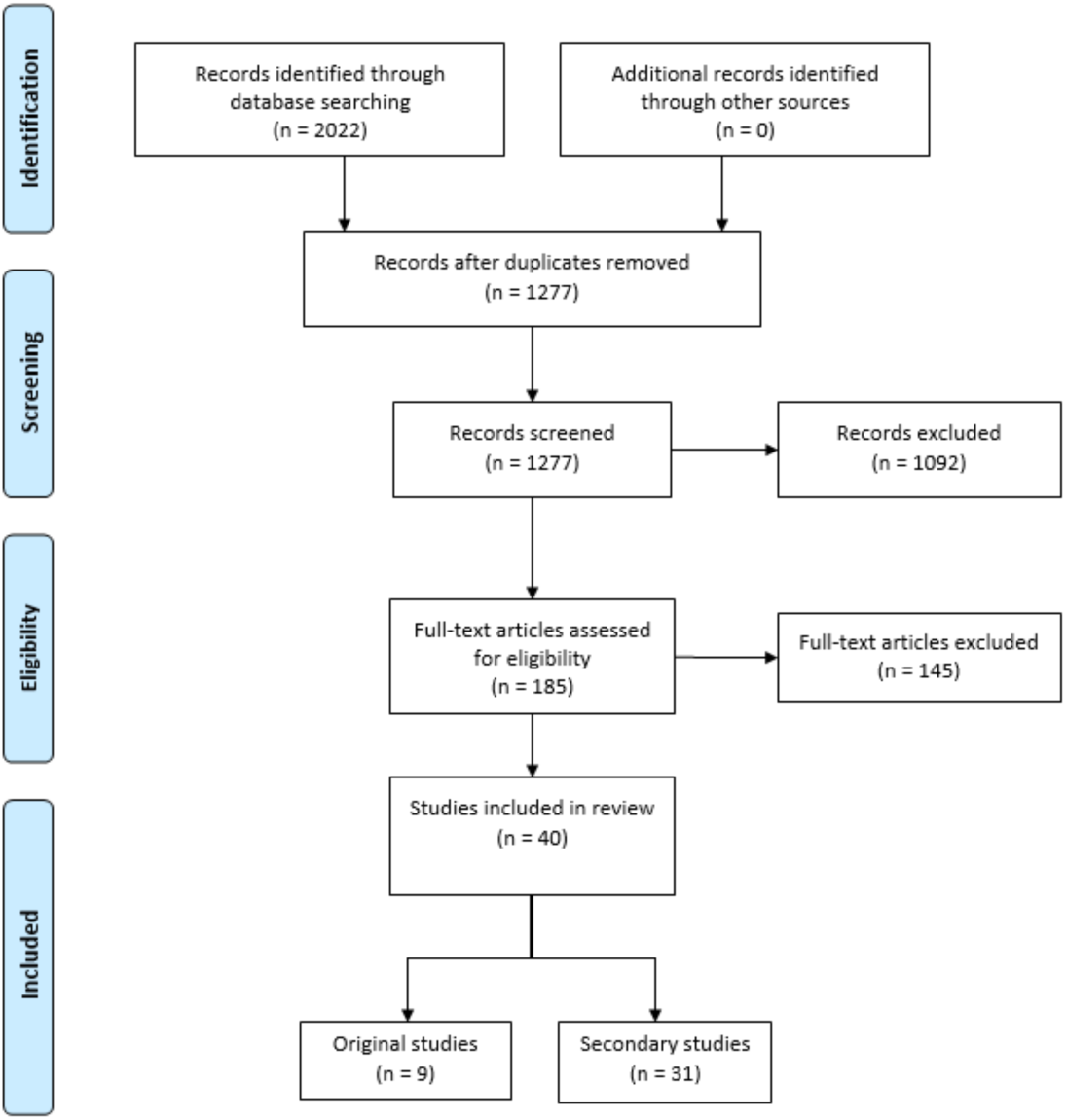
PRISMAFlow Diagram

## Results

Database searching identified 2022 potential articles; 1277 remained after duplicates were removed and from these 185 articles were subjected to full-text review. 145 articles were excluded for not meeting inclusion criteria, leaving 40 published studies included in the review.

Of the 40 articles, 9 were identified as original clinical trials; the remaining 31 articles are secondary investigations pertaining to the core trials, or follow-ups to original trials. 5 of the 9 studies used a controlled trial methodology. Of the 9 studies, 7 are completed and 2 are ongoing with published preliminary data. Of the completed trials, a total of 169 participants were enrolled, with 106 (70.2%) enrolled in controlled trials and 97 of these (64.2%) enrolled in trials pertaining to cancer-related anxiety and depression. Other presenting conditions studied include obsessive compulsive disorder, tobacco addiction, alcohol use disorder, treatment resistant depression and demoralization among long-term AIDS survivors.(Table 2.)

**Table 2.** Comparative table of key study characteristics of the results of eight psilocybin-assisted therapy trials. ^a^Study is ongoing. PSI = Preparation, Support, and Integration, CBT = Cognitive Behavioural Therapy, MET = Motivational Enhancement Therapy.

| Reference | Presenting Condition | # Subjects Recruited/ Usable Data/ Completed All Measures | Study Design | Type of Psychotherapy | Outcomes | Adverse Effects |
| --- | --- | --- | --- | --- | --- | --- |
| Moreno et al. (2006) | Obsessive - Compulsive Disorder | 9/9/9 | Modified Randomized Double Blind<sup>a</sup> | None | Acute reductions in OCD symptoms at 24 hours (>= 25% decrease in 8 patients, >=50% decrease in 6 patients) | Transient Hypertension (11%) |
| Grob et al. (2011) | Cancer-related anxiety and depression | 12/12/8 | Randomized, double-blind, crossover | None | Significant reductions in anxiety at 1- and 3-months follow-up and depression at 6 month follow up | None clinically significant |
| Johnson et al. (2014) | Tobacco addiction | 15/15/15 | Single Group, Open-Label | CBT | 12 of 15 participants demonstrated abstinence at 6 months follow up | Extreme or strong fear (40%), headache (53%), elevated BP and HR (NR) |
| Bogenschutz et al. (2015) | Alcohol use disorder | 10/9/9 | Single Group, Open-Label | MET | Percent drinking days decreased 1-7 weeks after psilocybin session. | Nausea and abdominal pain (1), Headaches (5), IBS-based diarrhea (1), insomnia following session (1) |
| Griffiths et al. (2016) | Cancer-related anxiety and depression | 56/51/46 | Randomized, double-blind, crossover | PSI | Significant decreases in depression and anxiety at 6 months follow-up | Headache (4%), nausea/vomiting (8%), physical discomfort (15%), psychological discomfort (22%), anxiety (21%), paranoia (2%) |
| Ross et al. (2016) | Cancer-related anxiety and depression | 29/29/22 | Randomized Double Blind Crossover | PSI | Significant and sustained anti-depressant and anxiolytic effects up 6.5 months follow-up | Headaches and migraines (28%), anxiety (14%), psychotic-like symptoms (7%), Elevated BP and HR (76%), nausea (14%) |
| Carhart-Harris et al. (2018) | Treatment resistant depression | 20/19/19 | Single Group, Open-Label | PSI | Maximal reduction in depressive symptoms for 5 weeks post-treatment with benefits persisting to 3 and 6 months | anxiety (79%), headaches (42%), nausea (26%), paranoia (16%) |
| Bogenschutz et al. (2018)<sup>a</sup> | Alcohol use disorder | 180/ n/a /3 | Randomized Double blind Parallel | CBT, MET | Self-reported decreasing craving and alcohol-associated problems, anxiety, and depression at final follow-up | Nausea and abdominal pain (33%) |
| Anderson et al. (2019)<sup>a</sup> | Demoralization due to AIDS diagnosis | 18/18/18 | Open-Label, Single Group | Group-based PSI | Rapid improvements in mood and anxiety symptoms | None clinically significant |

Of the 5 randomized control trials, 3 followed a crossover design, 1 was a parallel design, and 1 a modified escalating-dose design. In the crossover studies, each patient was exposed to both active placebo (niacin, diphenhydramine, or a very low dose of psilocybin) and an experimental dose of psilocybin in two sessions which varied between 1 and 7 weeks apart.

The trials themselves ranged in cohort sizes from 9 to 56 and the studies themselves lasted from 4 to 57 weeks (where reported). Each trial used a similar approach to psilocybin dosage and the setting for dose administration sessions. All trials used a synthetic formulation of psilocybin administered orally and largely in the medium to high dose ranges (25-40mg/70 kg).; dosage was calculated by one of three methods: mg, mg/k or mg/70kg.. The majority of trials administered psilocybin in multiple sessions (up to 3), separated by at least 1 week, and starting with lower doses (Table 3. Timeline of Psilocybin Trials). Sessions lasted up to 8 hours, and some trials provided overnight accommodation and all provided medical oversight and symptom management. Data was collected by various standardized tools and metrics including self-report, clinical assessment and biomarkers; one trial included brain imaging by fMRI.

**Table 3.**
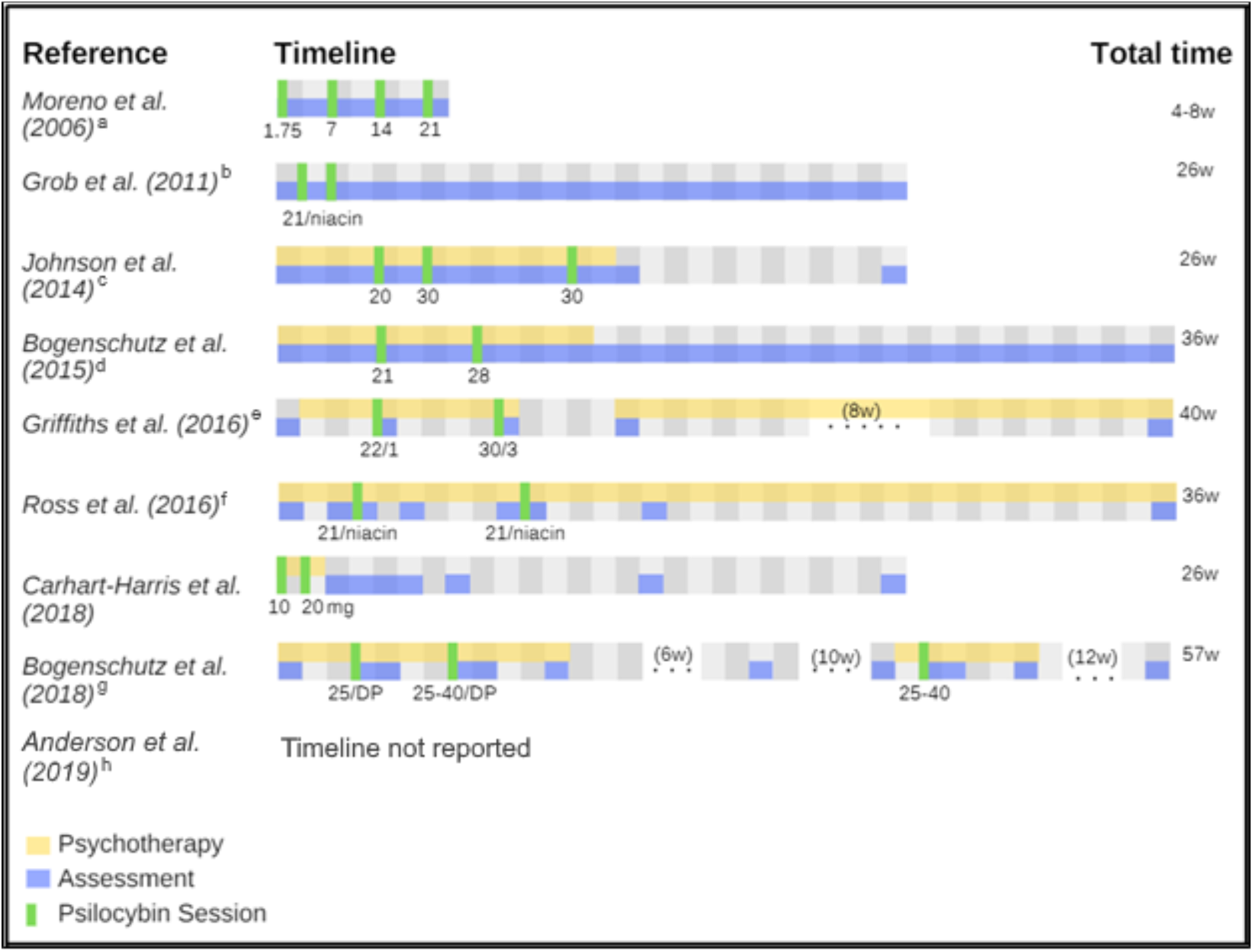
A comparative timeline of the eight reported psilocybin-assisted therapy trials. Each grid represents one week. All doses are in mg/70kg psilocybin except for Carhart-Harris et al. (2016) which is in mg. ^a^Doses progressively escalated between sessions. 1.75 mg/70kg placebo was randomly given in one of four sessions and time between sessions varied from 1-2 weeks. ^b^Participants received niacin placebo (250 mg) in one of two sessions. ^c^Participants given an optional third high-dose session. ^d^Original dose of 21 mg/70kg psilocybin was used in second session if participant preferred, there would be a significant risk, or the patient reported a complete mystical experience. ^e^Participants randomly assigned to low-dose session first (placebo-like) or high-dose session first, then crossed over for the following session. ^f^Participants allocated to control group first (niacin 250 mg) or experimental group first (21 mg/70kg psilocybin), then crossed over for the following session. ^g^Participants received either 25 mg/70 kg psilocybin vs. 50 mg diphenhydramine in the first session, and in the second session, depending on response, 30 or mg/70 kg psilocybin vs. 100 mg diphenhydramine. All participants were offered an additional third open-label psilocybin session from 25-40mg/70 kg. ^h^Patients were given four group-based preparatory sessions, one dosing session (21 mg/70 kg), followed by six group-based integration sessions.

**Table 4.**
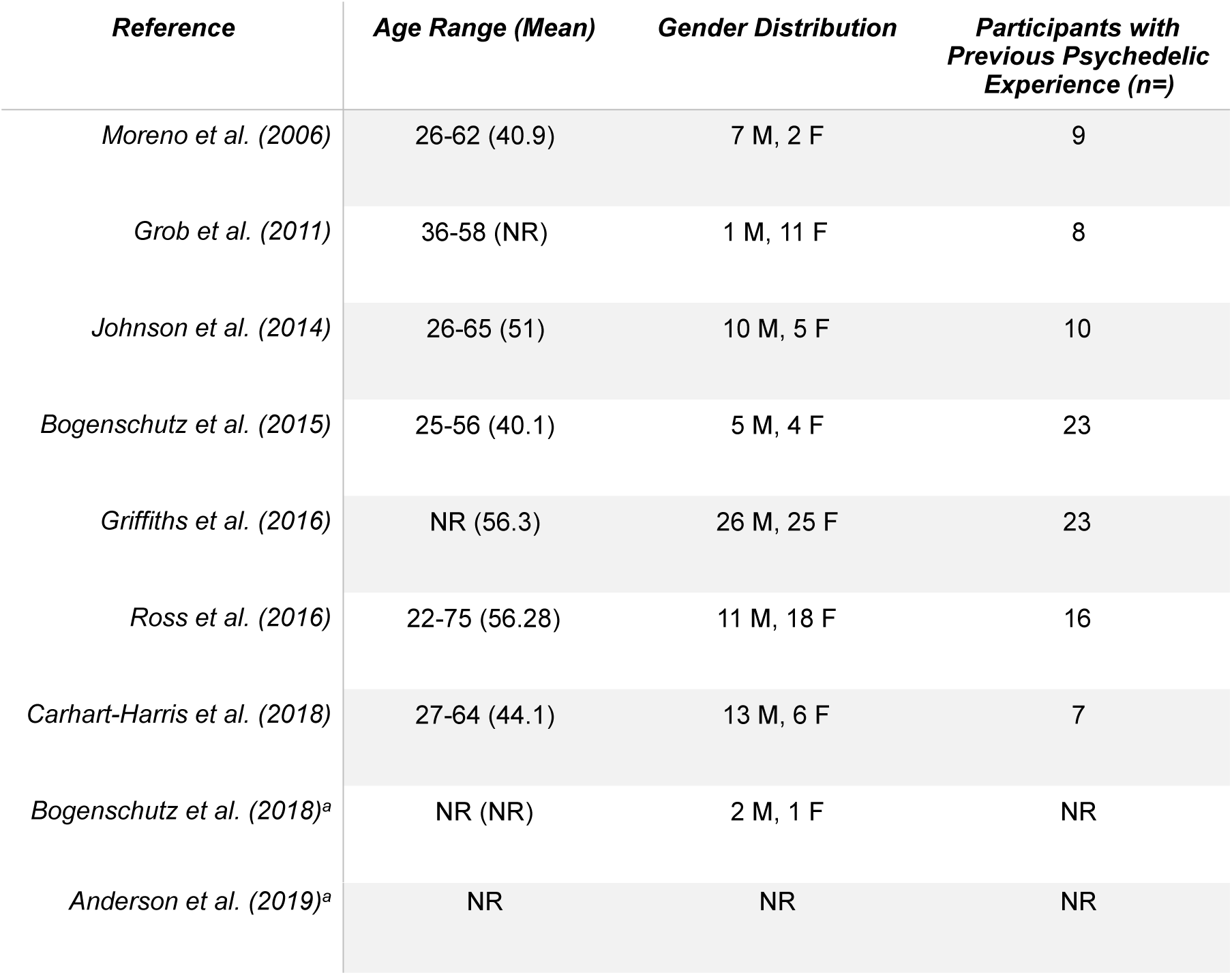
Comparative table of participant demographics of 9 psilocybin-assisted therapy trials. ^a^Study is ongoing

**Table 5.** Overview of ongoing psilocybin-assisted therapy trials. ^a^As of November 2^nd^ 2019

| **Principal Investigator and NCT** | **Presenting Condition** | **Status** | **Target Recruitment (n=)** | **Study Design** |
| --- | --- | --- | --- | --- |
| Woolley<br>(NCT02950467) | Depression from AIDS diagnosis | Active, not recruiting | 36 | Single Group, Open Label |
| Preller<br>(NCT03715127) | Depression | Recruiting | 60 | Randomized, Double Blind, Parallel |
| D'Souza<br>(NCT03554174) | Depression | Recruiting | 18 | Randomized Double Blind Crossover |
| D'Souza<br>(NCT03341689) | Migraine Headaches | Recruiting | 24 | Randomized Double Blind Crossover |
| Bogenschutz<br>(NCT02061293) | Alcohol Use Disorder | Recruiting | 180 | Randomized Double Blind Parallel |
| Raison<br>(NCT03866174) | Depression | Recruiting | 80 | Randomized, Double Blind, Parallel |
| Nutt (NCT03429075) | Depression | Recruiting | 50 | Randomized Double Blind, Parallel |
| Moreno<br>(NCT03300947) | Obsessive – Compulsive Disorder | Recruiting | 15 | Phase 1: Randomized Double Blind, Parallel<br>Phase 2: Randomized Single blind Parallel |
| Zisook, Worthy, Aaronson, Hellerstein, Soares, McIntyre, Licht, O'Keane, Schoevers, van der Wee, Somers, Bobo, McAllister-Williams, Young, Talbot<br>(NCT03775200) | Depression | Not yet recruiting | 216 | Randomized Double Blind Parallel |
| Griffiths<br>(NCT03181529) | Depression | Active, not recruiting | 24 | Randomized Single Blind Parallel |
| *Kelmendi*<br>(NCT03356483) | Obsessive -<br>Compulsive Disorder | Recruiting | 30 | Randomized Double<br>Blind Parallel |
| *Hendricks*<br>(NCT02037126) | Cocaine Use | Recruiting | 40 | Randomized Double<br>Blind Parallel |
| *Carhart-Harris, Nutt,*<br>*Rantamäki*<br>(NCT03380442) | Depression | Not yet recruiting | 60 | Randomized Double<br>Blind Parallel |
| *Johnson*<br>(NCT01943994) | Tobacco Addiction | Recruiting | 95 | Randomized Open-Label Parallel |
| *Griffiths*<br>(NCT04052568) | Anorexia Nervosa | Recruiting | 18 | Single Group, Open-Label |
| *Garcia-Romeu,*<br>*Rosenberg*<br>(NCT04123314) | Depression in<br>patients with<br>Alzheimer's Disease<br>or mild cognitive<br>impairment | Not yet recruiting | 20 | Single Group, Open-Label |

The majority of trials provided significant levels of therapy including preparation and integration, but also general psychological supports and, in the case of substance use disorder, additional therapies specific to addiction. Only the Grob (Grob et al., 2011) and Moreno (Moreno et al., 2006) trials did not report including psychotherapy or psychological supports. After varying degrees of preparation, trial subjects were given an oral dose of synthetic psilocybin in a quiet, protected and comfortable setting. Trials used curated musical playlists, eyeshades, and encouraged subjects to, in the words of one trial investigator “trust, let go, and be open” (Griffiths, et al., 2016). Where reported, trials included two guides (usually of differing genders), or sitters, who would accompany the subject through the sessions in an non-interventional, and supportive manner, later to debrief and help integrate the contents of the session. Overall, subjects tended to be middle aged, with mean ages between 40-53 where reported, and often had previous experience with psychedelics (Table 3).

In addition to the characteristics and outcomes reported in our summary tables, several reportable and common themes emerged from the literature. Those themes can be clustered into 5 distinct but related categorical statements.

### 1. Strength of the Subjective Psilocybin Experience Predicts Positive Treatment Outcomes

One finding consistent across several of the trials was that the intensity of psilocybin experience is strongly correlated with positive treatment outcomes. The acute subjective experience of the trial subject is the focus of many of the metrics employed, most commonly measured in Oceanic Boundlessness (OB), Dread of Ego Dissolution (DED) and Altered States of Consciousness scales. Many of the trial subjects report mystical-type experiences characterized by a deep sense of oneness and interconnectivity. Not all the trials found high rates of mystical experience however. In the Bogenshutz (2015) study on alcohol use disorder, only 3 of the 17 (17.6%) participants had *mysticomimetic* experiences, perhaps indicating a possible blunting effect from alcohol dependence. In general however, attributions of meaning or positive experience both correlated with improved clinical outcomes.

### 2. Psilocybin experiences are Rich, Highly Variable, Personalized and Memorable

All trials reported wide variation in experience among the subjects, but certain experiences were common to the phenomenology of psilocybin: awe, curiosity, connectedness, natural wonder, gratefulness, humility, oneness, compassion, love, acceptance, nature relatedness, optimism, forgiveness and acceptance were commonly referenced, and subjects generally rated the experience as intense, memorable while program staff observed wide variation of experience, with each unique to the history, personality, and life situation of the subject. Positive attribution towards the psilocybin session was found to remain high upon long-term follow up.

### 3. Transient Psychological Distress During Psilocybin Sessions is Common and Normative as are Post-session Headaches

No serious or life-threatening adverse effects were experienced in any of the trials significant enough to question the safety of psilocybin in a controlled setting. However, transient side effects during psilocybin sessions, often experienced as challenging, were reported: acute reactions of confusion, fear, paranoia, anxiety, nausea, headache, disorientation and transient psychological distress were reported in as much as 40% of trial subjects were reported across several trials. Significantly, several trials found post-treatment headaches to be common in 40-50% of subjects. (Grob, et al., 2011, Carhart-Harris, et al, 2016); a pattern of dose-dependent, delayed and transient headaches has been documented in healthy volunteers administered psilocybin (Johnson et al., 2011).

### 4. Psilocybin May Alter Characteristics of Personality

Experiences under psilocybin tended to give participants lasting, sustained insights leading to shifts in self-identity. Psilocybin trial participants experienced alterations to traits of personality— increased openness, extraversion and conscientiousness, decreased authoritarian political views. Traits of empathy, openness, optimism and conscientiousness were markedly improved; in contrast anhedonia, pessimism, fear and authoritarian political views were all found to decrease. Many subjects experienced decrease neuroticism, and a increases in acceptance resulted in experiences of forgiveness, release or acceptance of one’s life.

### 5. Psilocybin is an Experience of Meaning-Making

Many trial subjects had experiences under psilocybin that remained significant and meaningful when measured for up to 18 months. Many reported being able to see problems from new perspectives, and to see how they could change the narrative of their lives. Many reported being able to see their lives from a bigger, and more supported, perspective. The fear of death, for many, was lessened by insight into their place in life, and of the experience of a spirit or presence not dependent on the body. Many subjects reported their psilocybin experiences as among the top 5 meaningful experiences of their lives.

### 6. Program Variables and Psychological Supports Play a Significant Role in Treatment Outcomes

Subjects were asked about their response to the music playlists which accompany the dosing sessions, with feedback on both favourable and unfavourable experiences. Music was found to help guide a session, and could heighten emotional response, but many subjects reported dissatisfaction with either the choice, or timing of music played. Subjects were found to have a generally good rapport with program staff, and many cited their relationships with program staff as important to their experience. Sensitivity to setting is a characteristic of the psilocybin experience, and trial protocols ensured a comfortable, relaxed setting.

What emerges from the scoping review is a new but relatively consistently applied experientially-oriented psycho-pharmacotherapy, best combined with psychological supports and conducted in a thoughtful, appropriate setting. A relative consensus emerges of a medium to high dose protocol, with an escalation of dosage over 2-3 treatment sessions, prefaced by intensive psychological preparation and post-treatment integration.

## Discussion

Trials reported several limitations which significantly limit the generalizability of findings. Several of the trials were not controlled, but rather open label exploratory trials to determine safety and efficacy. In addition, several of the trials included a significant number of widespread metrics and multiple assessment tools, indicating a lack of specificity in identifying mechanisms of action. Consistently and despite various approaches, trials reported a strong difficulty in truly blinding and staff due to the obvious effects of psilocybin. Even the lower doses of psilocybin were found impossible to blind.

Cancer trials showed the highest level of methodological rigour, as each used a controlled crossover design. The nature of the crossover design in the included clinical trials led to reported unblinding of both participant and investigator, allowing for influencing of expectations (Grob et al., 2011; Griffiths et al., 2016, Ross et al., 2016) and limited the assessment of efficacy and clinical benefit after the crossing over of treatments (Griffiths et al., 2016; Ross et al., 2016). Crossover designs which employ a placebo also suffer from the potential of carryover effects for the experiment-first, placebo-second group. This can be minimized through a wash-out period, which varied between one to seven weeks for the three studies (Grob et al., 2011; Griffiths et al., 2016, Ross et al., 2016).

Trials also tended to have very small subject sample sizes. The relatively homogenous patient demographics limit the generalizability of the psilocybin trial findings. It is not clear if this modality would be effective with patients characterized by greater mental health co-morbidity, marginalization, poverty, instability or relative youth. Trials reported common exclusion criteria: personal or family history of schizophrenia/other psychotic disorders or bipolar disorders, personal cocaine, psycho-stimulant or opioid dependence, or family history of suicide. RCTs will remain challenging due to the difficulty in blinding study participants, and because of the confounding variable of the psychotherapies provided.

Psychological therapies given in addition to the psilocybin session confound the study outcomes, if what we are assessing is the therapeutic potential of psilocybin. Further, it must be asked if the peak psychedelic model is the only viable practice model to investigate; regimens of lower doses, plus collective group models, remain to be studied. Psilocybin at microdoses also requires more rigorous investigation. A recent systematic study found microdosing improves mood, energy levels and cognition, creativity, optimism and openness, as well as alters perceptions of time (Polito & Stevenson, 2019) and a recent self-report study of microdosers found reductions in depression, stress and distractibility with increased absorption and neuroticism (Anderson, et al., 2019). Smaller sub-perceptual doses would allow for more relaxed trial inclusion criteria, and could be possibly be used in the treatment of PTSD given the role of low-dose psilocybin in reducing fear response in laboratory animals (Catlow et al., 2013).

The ayahausca church model has been associated with lower rates of substance use disorder and overall improvements in health. (McKenna, 2004, Barbosa, Mizumoto, Bogenschutz & Strassman, 2012, Fabregas et al. 2010) while ceremonial peyote rituals have been proposed as a model of *ethnopharmacologic* treatment for alcohol and other forms of substance use disorder (Prue, 2013, Blum et al. 1977). Collective ritual ceremony has the added benefits of socialization, peer support and structure, all factors thought to be beneficial in the process of recovery and all characteristic of the 12-step recovery model. Further, while much has been made of the potential for psilocybin to induce mystical-type experiences (Griffiths et al. 2006, 2008), studies have found this to be not always be the norm; Cummins and Lyke (2013) found 47% of naturalistic users to report peak experiences while under psilocybin. Subjects reported a range of powerful experiences in these trials; meaningful and often challenging but not always mystical. A more sophisticated mapping of consciousness and understanding of altered states remain necessary.

An assessment of registered NCT trials (Table 3.) identified a further 16 new clinical trials active, recruiting or planned in the U.S. and Europe. In a notable shift, 9 of these new trials follow randomized, double blind parallel designs. Parallel designs give the advantage of avoiding carryover effects. They are also favoured in larger studies; the newer trials are trending to be larger than those reviewed here; they range in target sample size from 18-216 (mean, 66). One currently recruiting trial uses psilocybin in the treatment of headaches, building on earlier indications that chronic migraine sufferers or those with cluster headaches report relief via self-medication with psilocybin as well as LSD (Sewell et al., 2006); another recruiting trial for the treatment of cocaine use disorder is the first of it’s kind.

Popular interest in the potential benefit of serotonergic psychedelics will necessitate new models of policy, regulation and practice. Haden (Haden et al. 2016) has proposed a public health model of oversight including the regulation of a new classification of psychedelic therapists. Training programs, certification and manuals remain to be developed. The current prohibition of psilocybin (as well as other classic psychedelics) remains a barrier to science: researchers must currently apply for difficult and time-consuming special access and many countries may lack domestic suppliers. Following cannabis, the decriminalization patterns newly evidenced may evolve more quickly than the scientific research, necessitating the rapid development of evidence-based practice models and indications for use while we continue to investigate the potential clinical benefits by rigorous scientific method. The therapeutic potential of *psilocybe* mushrooms may not be limited to the psilocybin compound alone; psilocin, baeocystin, norbaeocystin, aerguinascin and norpsilocin are also found in *psilocybe* mushrooms and may themselves have value alone or in entourage.

## Conclusion

The psilocybin trials have demonstrated safety, tolerability and efficacy in treating a range of mood and self-regulatory disorders, indicating a common underlying basis to distinct pathologies. Outcomes are positive both in the studied reductions of problematic behaviours, but also in the sustained elevation of mood. Adverse effects are rare, though common side effects include transient anxiety, nausea and post-treatment headaches.The psilocybin trials have established a marked ability to interrupt disordered psycho-pathological processes and to lead to improvements in health behaviours. The mechanism of action is not yet fully understood. Current legal prohibition on psilocybin hinders basic science and continued clinical investigation.

The use of psychedelics represents a paradigm shift in our approach to disorders of mental health (Nichols, et al., 2017, Schenbert 2018), including addiction (Morgan et al., 2017). The promise of psilocybin therapy rests in it’s apparent ability to disrupt dysphoric mood states and obsessive or compulsive self-regulatory disorders. More rigorous scientific study is necessary to lay the basis for evidence-informed practice.

## Data Availability

Supplemental Information posted with article. Corresponding author may be reached at email provided for further information.

